# Beyond proportional recovery in wake-up stroke: unsupervised recovery clusters based on the NIHSS

**DOI:** 10.1101/2025.06.04.25328943

**Authors:** Andrea Zanola, Antonio Luigi Bisogno, Veronika Vadinova, Götz Thomalla, Bastian Cheng, Manfredo Atzori, Maurizio Corbetta

## Abstract

Post-stroke rehabilitation is a complex process influenced by several neurophysiological factors. The recovery is traditionally predicted based on initial impairment using linear models. The Proportional Recovery Rule (PRR), developed on the Fugl-Meyer scale, has even been proposed as a therapeutic target. In this framework, patients are classified as “fitters” or “non-fitters”, though this distinction depends on the methodology used. Additionally, issues like mathematical coupling and ceiling effects on clinical scales could raise concerns about the validity of these models. To overcome these issues, Repeated Spectral Clustering (RSC) was used to identify recovery patterns based on NIHSS. We selected 201 patients from the WAKE-UP trail, all moderately impaired at onset and still impaired at 22/36 hours. Clustering was performed using a similarity matrix based on pairwise absolute differences between recovery ratios, calculated from 22/36 hours to 90 days post-stroke. Cluster differences were tested with prognostic factors, including lesion volume, side, treatment, and the Heidelberg scale. The PRR was fit to the cohort for comparison with clustering results. The linear fit reproduced findings consistent with the literature, such as a correlation of ***ρ*(*x*, Δ) =** .**73** and an average recovery ratio of 70% for the “fitters”. RSC grouped patients into six recovery clusters: ***C***_**0**_ (full recovery), ***C***_**1**_ (above average), ***C***_**2**_ and ***C***_**3**_ (average, PRR-aligned), ***C***_**4**_ (below average), and ***C***_**5**_ (deterioration). NIHSS scores in most patients declined non-proportionally. Lesion volume was not significantly different across clusters, while left-sided strokes were higher in low recovery clusters. Patients with a recovery ratio **≥ 0.3** within two weeks mostly fell into favorable clusters (***C***_**0**_–***C***_**3**_), covering **≈ 90%** of such cases. The identified clusters provide a refined view of stroke recovery following wake-up stroke. Clustering better captures patient similarities, enabling the assessment of neurophysiological differences between groups and supporting tailored interventions.

## 1 Introduction

Recovery after stroke is a complex and multifaceted process that involves the interplay of neurological, physiological, and environmental factors. Longitudinal datasets offer a valuable opportunity to investigate recovery trajectories over time to identify prognostic factors and therapeutic targets. Traditional approaches to studying stroke recovery have relied heavily on evaluating outcomes in acute versus chronic phases. Although these methods yield important insights, they often fall short in capturing the heterogeneity of recovery patterns between individuals. A key concept in this domain is the categorization of patients into “fitters” vs. “non-fitters”, based on how closely their recovery trajectories align with established mathematical models [1, 2].

However, this dichotomy and related framework have notable limitations. First, from a methodological perspective, some authors [3, 4], have raised concerns about the issue of mathematical coupling, since proportional recovery is calculated based on acute vs. acute minus chronic scores, which inherently links the two variables. Second, widely described in recent literature [5], the ceiling effect is present in commonly used clinical scales. Essentially, this reflects a reduced sensitivity to the variability currently observed, implying that there may be additional variability that remains undetected. This leads to the possibility that unexplained variability is underestimated [3]. Another major concern, from a physiological standpoint, is that this theoretical framework has been interpreted to suggest that normal recovery entails regaining approximately 70% of the initial deficit. Such an interpretation risks oversimplifying the complex and individualized mechanisms of stroke recovery. Most importantly, this proportional recovery has often been interpreted as ground truth and, in certain cases, can be considered a valuable therapeutic target to obtain [6]. Unfortunately, strong neurophysiological evidence in this regard is still lacking, and most authors have focused on specific deficits and clinical scales (i.e. Fugl Meyer [7] and motor deficits). The literature reflects a lively debate, with some studies arguing for the utility of the “fitters” vs. “non-fitters” categorization and others highlighting its constraints and potential biases [4, 8–10].

In light of these challenges, the present study aims to provide a complementary approach to describe the recovery of stroke patients by using unsupervised techniques. Classical methodologies are focused on the statistical evaluation of the goodness-of-fit of a model for the whole population, assessing how one or more independent variables describe one or more dependent ones (e.g., how acute score describes the chronic one). Clustering, one of the main unsupervised techniques, is particularly effective in finding groups of subjects, i.e. phenotypes, within the analyzed population. Specifically, while statistical techniques focus primarily on correlation between variables, clustering instead focuses on grouping subjects based on their similarity to one another. From this perspective, clustering can be seen as an analysis of the “subject correlation” representing a complementary, but opposite, approach to the conventional analysis of feature correlation [11, 12]. As mentioned in Yang *et al*. [13], clustering is one of the most useful methods for analyzing similarities between patients for precision medicine. It groups patients into subsets that are clinically meaningful and can be used for a variety of tasks, such as policy making and personalized therapies. Kim *et al*. [14] stresses that finding similar clusters among stroke patients can be helpful from a medical perspective, as it can lead to the discovery of new patterns and ways to manage stroke more effectively.

In this work, we apply clustering algorithms tailored for biomedical data analysis [11] to the recovery ratio (*RR*) [15, 16] of patients, based on the National Institute of Health Stroke Scale (NIHSS) [17], in a large longitudinal sample of wake-up [18] stroke patients. The recovery ratio used for clustering is calculated using the total NIHSS scores measured in the subacute phase (22/36 hours) to 90 days. This approach presents several advantages over the existing methods. Clustering allows for the identification of diverse recovery patterns without presupposing specific models, in a data-driven manner. Moreover, it helps mitigate the issue of mathematical coupling inherent in traditional proportional recovery calculations by avoiding any model fitting in the statistical sense.

## 2 Methods

### 2.1 Dataset

WAKE-UP [18] is an investigator-initiated, multicenter, randomized, double-blind, placebo-controlled clinical trial involving stroke patients with an unknown time of onset. Patients were randomly assigned to receive intravenous alteplase (rtPA) or placebo. The study was registered under the identifier NCT01525290. The dataset includes 503 patients, with NIHSS [17] scores measured at four time points: onset, 22/36 hours, 7 days, and 90 days.

For the purposes of this study, patients with a Level-of-Consciousness Vigilance (LOC-V) score of zero and complete NIHSS data at all four time points (no missing values) were selected. Patients were further selected based on stroke severity, as measured by the total NIHSS score at onset. Only patients with a total NIHSS score greater than 4 (moderate stroke severity) were included; patients with a total NIHSS score of zero at 22/36 hours after the event were further excluded from the analysis. These selection criteria were designed to reduce the inherent ceiling effect of the NIHSS and focus on patients with persistent deficits. As a result, the final analysis focused on 201 patients who were still impaired at 22/36 hours. In the supplementary material Section 1 a summary schema of the inclusion criteria and the consequent effect on the size of the population is reported.

For each subject, general demographic information (age, sex and treatment group), lesion characteristics (lesion side, lesion volume, large vessel occlusion and lacunes), the 90-day modified Rankin Scale (mRS) [19], the Heidelberg hemorrhage classification [20], and the NIHSS with all 15 sub-items are available. The mRS scale assesses disability and dependence after a stroke, while the Heidelberg classification categorizes intracranial hemorrhages following ischemic stroke. The demographic characteristics of the selected patients are summarized in Table 1. A *χ*^2^ independence test [21] did not reveal statistically significant differences in the distribution of male and female participants between the treated and non-treated groups (*χ*^2^(1, 201) = 0.44, *p* = 0.51). Moreover, there are no significant differences in terms of median age between the four groups, as assessed by the Kruskal-Willis test [22] (*H*(3) = 3.53, *p* = .32). For this work, a significance level of *α* = 0.01 was chosen.

**Table 1.** Demographical variables. The table reports the number of subjects divided by the two attributes, group (rtPA treated vs. non-treated) and sex (female vs. male). It also reports the mean ages for each group, with the standard deviation in years.

| Group \ Sex | Female | Male |  |
| --- | --- | --- | --- |
| rtPA treated | 36 (63.6 ± 12.1 years) | 58 (64.8 ± 10.5 years) | 94 |
| non-treated | 47 (67.5 ± 10.5 years) | 60 (65.4 ± 11.6 years) | 107 |
|  | 83 | 118 | 201 |

Regarding the lesion side, among the 201 subjects, 121 (60%) had left side lesions, 75 (37%) had right side lesions, and 5 (3%) had bilateral lesions. The average lesion volume, measured at hospital admission using apparent diffusion coefficient MRI maps, was 8.3± 12.1 mL (mean ± standard deviation). The distribution of cardiovascular risk factors for the 201 selected subjects is reported in Table 2.

**Table 2.** The cardiovascular risk factors of the 201 subjects selected for this study are presented. Values represent the number of subjects.

| Cardiovascular Risk Factor | Non-Present | Present | Missing Value |
| --- | --- | --- | --- |
| Arterial Hypertension | 102 (51%) | 98 (49%) | 1 |
| Atrial Fibrillation | 168 (84%) | 30 (15%) | 3 |
| Transient Ischemic Attack | 192 (96%) | 5 (2%) | 4 |
| Previous Ischemic Stroke | 176 (88%) | 25 (12%) | 0 |
| Hypercholesterolemia | 121 (62%) | 74 (38%) | 6 |
| Diabetes mellitus type II | 162 (82%) | 36 (18%) | 3 |

### 2.2 Fitters versus non-fitters calculation

The chronic and sub-acute scores can be plotted against each other, as shown in Figure 1, Panel A. However, the most commonly used plot in the literature is shown in Panel B, which highlights the ceiling effect [4], indicated by the green line. The ceiling effect occurs when data points reach the maximum limit possible on the measurement scale, i.e here 100% recovery. The correlation between the two variables, measured using the Spearman correlation, is strong and highly significant (*ρ*_*s*_(201) = .73, *p <* .001). This value is consistent with similar findings reported in the literature [23]. A linear model can be used to fit the change in scores (sub-acute score minus chronic score) as the dependent variable, with the sub-acute score as the independent variable. In this work, the linear model is fitted without the intercept; the medical and statistical reasons behind can be found in the supplementary material Section 2; the proportional recovery rule (PRR) is shown in Figure 1, Panel B, with a dashed black line. In both panels of Figure 1, the 95% confidence interval (CI), representing the uncertainty of the fitted model, is shown in gray; the 70% prediction interval (PI) instead, which estimates where future observations are expected to fall, is shown in light blue. Both intervals were calculated following the approach described by Howell [24], 2007; for further details, see the supplementary material Section 3. The prediction interval can be used to categorize the population into “fitters” and “non-fitters”. In both panels A and B of Figure 1, “fitters” are marked with a circle (∘), while “non-fitters” with a cross (×). The prediction interval contains 70% of the subjects, who show an average recovery ratio of 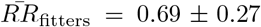. This is in line with the well-established literature on the proportional recovery rule in stroke [23].

**Fig. 1.**
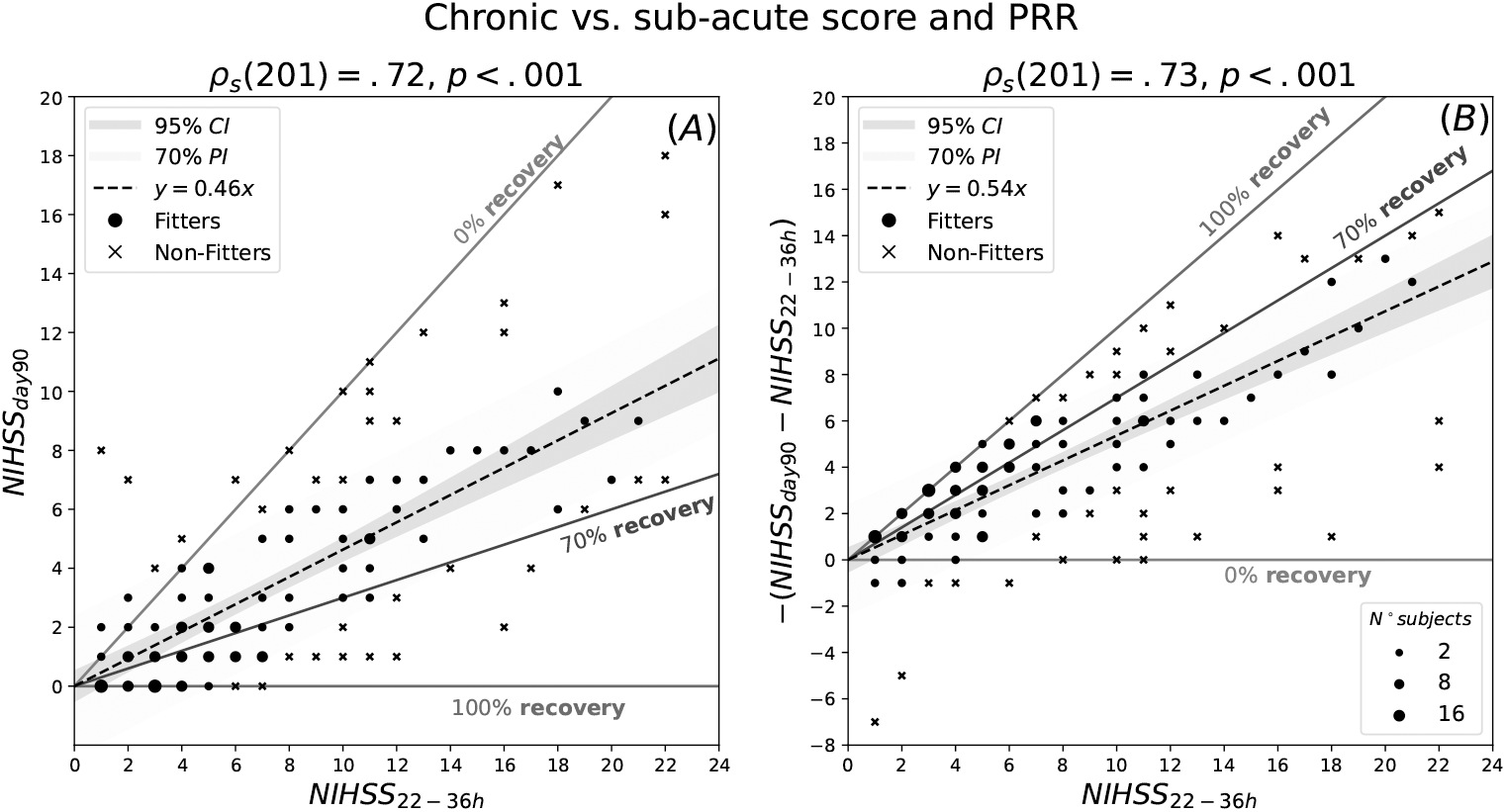
Chronic vs. sub-acute score and PRR. Panel A, shows the total NIHSS at 90 days vs. total NIHSS at 22/36 hours (chronic vs. sub-acute). Panel B, shows the PRR i.e. the total NIHSS change vs. total NIHSS at 22/36 hours (change vs. sub-acute). In both panels, the title reports the Spearman’s correlation. In both panels the red, blue and green lines indicate 0%, 70% and 100% recovery respectively. The dashed line represents the fitted linear model (PRR), in grey the 95% confidence interval (CI) and in light blue the 70% (≈1*σ*) prediction interval (PI). Subjects in both panels are marked with a circle (∘) and identified as “fitters” if they stay within the prediction interval (light blue band), and with a cross (×) otherwise (“non-fitters”). The size of the markers, represent the number of subjects having that combination of values, as indicated by the corresponding legend.

However, the distinction between “fitters” vs. “non-fitters” (or outliers) is far from being unique. Different techniques dichotomize the population in different ways, high-lighting different aspects of the data. An notable effort has been made to compare different techniques in Kundert *et al*. [5], 2019. In order to briefly review and compare these methods, they have been applied to the population under study. Seven methods to define “non-fitters” or outliers have been used and reported in Figure 2; methodological details can be found in the supplementary material Section 4. The distribution of the recovery ratio for the entire population is also reported in the same figure. Interestingly, this distribution does not follow a normal distribution centered around 70%.

**Fig. 2.**
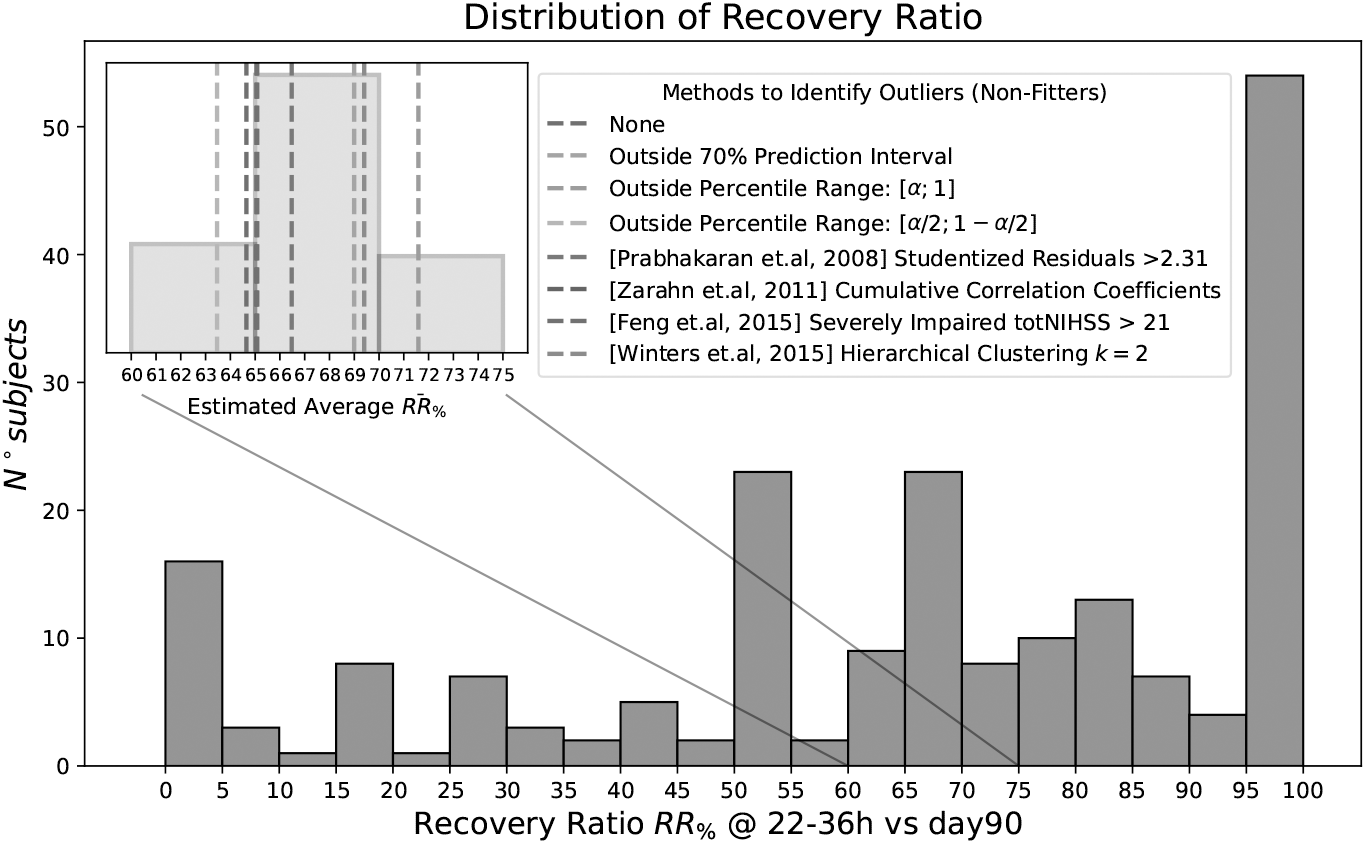
Recovery ratio distribution. The main histogram depicts, in gray, the distribution of recovery ratios for the entire population. The inset panel provides a magnified view focusing on the recovery ratio range between 60% and 75%, highlighting the average recovery ratio estimates obtained from various methods. Different colors correspond to different estimation approaches, as indicated in the legend. For methodological details, refer to the supplementary material Section 4.

The variability in the average recovery ratios arises from the different criteria and algorithms applied between methods. Classifying the same patient as a “fitter” or a “non-fitter” depending on the method used, highlights the inherent limitations of an arbitrary dichotomization. In addition, Figure 2 clearly shows that the proportional recovery rule does not represent the typical amount of recovery (i.e. the recovery ratio is not normally distributed). This work aims to move beyond this classification and advocates for a more nuanced, data-driven approach: clustering.

### 2.3 Recovery Ratio

A recovery ratio index (*RR*) of the form (acute score - chronic score)/acute score, was used to study the degree of recovery, as described in previous work [15], [16]. Let’s denote as *S*, a score or a measure of choice, aimed at quantifying the impairment; then the recovery ratio index is defined as

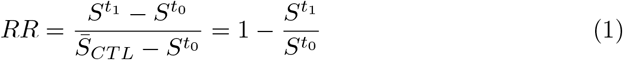

In Equation 1, 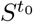 represents the sub-acute score measured at an initial time point *t*_0_, while 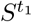 represents the chronic score measured at a later time point *t*_1_. The term 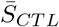denotes the average score for the control population. When considering the total NIHSS as the score *S*, 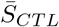 equals zero, since healthy individuals have a total NIHSS of zero, which simplifies the formula. This formula applies only when the sub-acute score is greater than zero, i.e., only for impaired patients. Since some patients exhibit a worse condition at the chronic time point than at the sub-acute stage (deterioration), the recovery ratio can be negative in these cases. For the selected sample of 201 subjects, the average recovery ratio is 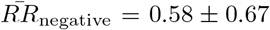. Since the recovery ratio is intended to range between 0 and 1, the negative values are set to zero. With this convention, the average recovery ratio becomes 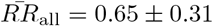.

### 2.4 Clustering

Clustering is a popular unsupervised technique, particularly useful in the identification of phenotypes [14], which can be effectively used to find groups of patients with common recovery. The following procedure was applied to cluster the data according to the recovery ratio.

1. The total NIHSS score at 22/36 hours (sub-acute score) and the total NIHSS score at 90 days (chronic score) are used to calculate the recovery ratio (*RR*) for each subject, as defined in Equation 1.
2. The distance between subjects is calculated using the Manhattan distance on their recovery ratios. Between two subjects *i* and *j*, the distance is *d*_*ij*_ = |*RR*_*i*_ −*RR*_*j*_|. Distances are divided with respect to the maximum value found in the dataset.
3. The matrix *D*_(*ij*)_ = *d*_*ij*_ is obtained from the pairwise distances between patients. After that, the similarity matrix (*W* = I − *D*) is calculated and used as input for the clustering algorithm.
4. The application of the Repeated Spectral Clustering (RSC) [11] algorithm to the similarity matrix *W* allows the identification of groups of patients with similar recovery ratios.

The RSC algorithm is a blend of spectral clustering [25], known to work well with similarities matrices representing graphs, and consensus clustering [26], known to be powerful since it allows to deal with the variability of results that stem from random initializations. In brief, RSC takes the matrix *W* as input to the spectral clustering procedure. Instead of running *k*-means just once, it is performed *N* times to account for randomness in centroid initialization. Across these *N* iterations, the number of times two patients are assigned to the same cluster is recorded. The more frequently two subjects are clustered together, the stronger the evidence that they should remain in the same cluster in the final assignment. The final cluster assignment is based on the co-occurrence matrix *C*, where each element *C*_*ij*_ represents the number of times subjects *i* and *j* were clustered together across all runs. For a complete description of the algorithm, including technical details and its application to NIHSS scores, refer to Tshimanga *et al*. [11], 2025.

The optimal number of clusters is determined by calculating the maximum spectral gap for different cluster configurations (i.e. different *k*) [11]. The elbow criterion is a simple and effective way to find the optimal number of clusters. In particular, for *k* = 6, the maximum spectral gap is at the end of a high plateau, indicating well-separated groups, while for *k* = 7, it decreases sharply, indicating subjects that are worst separated in groups. Thus, *k* = 6 is chosen as the optimal number of clusters. For more details, see the supplementary material Section 5.

The clustering algorithm uses the recovery ratio, which is derived from the total NIHSS at two distinct time points. The total NIHSS constitutes the aggregate of the 15 individual items that comprise the scale. All other features, including general subject information, lesion information, 90-day mRS, and the Heidelberg hemorrhage classification, were excluded from the clustering process. Instead, these features were used as validation features to characterize the resulting clusters and assess differences between them. The numerical and ordinal features between the six groups have been compared with the Kruskal-Wallis [22] test as omnibus, and then the Mann-Whiteny U [27] as post hoc for pairwise comparisons. Boolean or categorical features have been compared between the six groups with the *χ*^2^ independence test as omnibus and post hoc for pairwise comparisons. The p-values of the omnibus tests, as well as those of the post hoc test, were corrected for multiple comparisons using the FDR Benjamini-Hochberg correction [28]. The statistics used in this work to evaluate the effect size of the aforementioned tests are reported in the supplementary material, Section 6.

### 2.5 Chronic disability and early recovery

The early recovery ratio, calculated between 22/36 hours and 7 days as described in Equation 1, was also considered in this study. The 90-day mRS was dichotomized into two groups: scores ≤ 2, and scores greater than 2. This threshold is clinically relevant, as an mRS score of 2 indicates a mild disability that does not interfere with daily activities. To examine the relationship between early recovery and long-term outcomes, we compared the number of subjects with mRS scores greater than 2 versus those with scores of 2 or less across different recovery intervals. The 95% confidence intervals for these proportions were estimated using binomial statistics and are shown in Figure 5, Panel A. Additionally, the percentage of subjects in each mRS group across recovery intervals is presented in Figure 5, Panel B.

## 3. Results

### 3.1 Clustering Recovery Ratio

The repeated spectral clustering (RSC) algorithm, as described in section 2, uses exclusively the recovery ratio defined in Equation 1. This variable is reported against the total NIHSS at 22/36 hours (sub-acute) in Panel A of Figure 3. Although clustering is performed on the similarity matrix *W*, as described in subsection 2.4, the visible effect of the algorithm is horizontal separation between subjects. This allows to have clusters with a defined recovery ratio with low variation. For example, cluster *C*_0_, which contains all subjects with a perfect recovery ratio of 100%, has a null standard deviation; *C*_5_ instead, is the cluster with the worst mean recovery ratio which is close to 1%. Within this group, there are all subjects with a chronic score worse than sub-acute one. At the population level, the sub-acute score is significantly but moderately associated with the recovery ratio (*ρ*_*s*_(201) = −.34, *p <* .001).

**Fig. 3.**
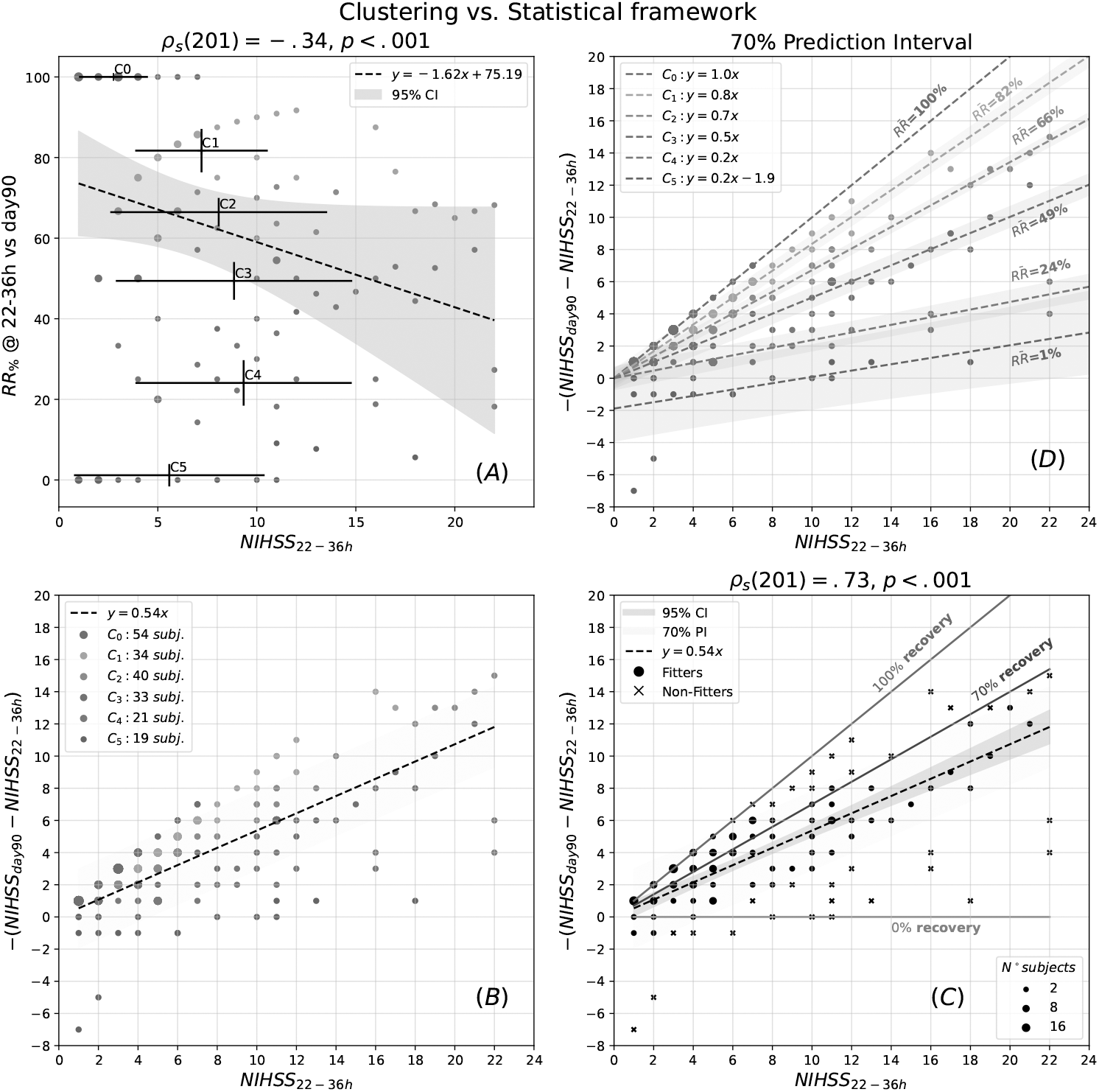
Clustering results. Panel A, shows the recovery ratio (RR) vs. the total NIHSS at 22-36h, with colors indicating different clusters. Error-bars are ported for both variables and for each cluster. In gray is reported the 95% confidence interval (CI). Panel B and C are shown in order to compare how patients are colored based on cluster results vs. how they are labeled as “fitters” and “non-fitters”. Panel C is equal to Panel B in Figure 1, and visualized here for convenience. Finally Panel D, shows the six fitted linear models (proportional recovery rules) for the subjects contained in each clusters. The median recovery ratio for each cluster and the 70% prediction interval (PI) for every model is reported as well. The size of the markers in each of the panels represents the number of subjects having that combination of values, as indicated by the corresponding legend in Panel C.

The bottom part of Figure 3 illustrates the difference between how patients are grouped using a clustering approach (Panel B) versus how they are divided in “fitters” and “non-fitters” using the traditional statistical method (Panel C). Interestingly, cluster *C*_3_ closely follows the fitted linear model, shown by the black dashed line.

Panel D of Figure 3 shows the proportional recovery rules obtained fitting the subjects for each cluster. As reported in the legend, there is a strict correspondence between the average *RR* of each cluster and the angular coefficients of these lines. The 70% prediction interval is reported for every fitted line. Looking at the prediction intervals of each cluster, these models discriminate subjects at one standard deviation, when the sub-acute NIHSS score is greater than six. The cluster *C*_5_ (in brown) overlaps with the cluster *C*_4_ (in violet), probably due to the few subjects assigned to the first (19), and consequently a broad prediction interval. Moreover, cluster *C*_5_ is clearly associated with people who actually become more impaired at 90 days compared to 22/36 hours (the intercept is −1.9). Only for the cluster *C*_5_, a linear model with the intercept is significantly better than one without (*F* (1, 17) = 9.08, *p <* .01, *f* ^2^ = 0.53). \

In conclusions, while the PRR gives a methodological way to identify “fitters” vs. “non-fitters”, the clustering-based procedure gives rise to a data-driven family of groups, sharing similar recovery. As a consequence, subjects are not distinguished anymore between “fitters” vs. “non-fitters”, but rather between levels of recovery. In summary, *C*_0_ (blue) has perfect recovery, *C*_1_ (orange) has above average recovery, *C*_2_ (green) and *C*_3_ (red) have average recovery (in line with the expectations of the PRR), *C*_4_ (violet) has below average recovery, and finally *C*_5_ (brown) has no recovery.

### 3.2 Differences between Clusters

The clustering algorithm uses only the recovery ratio, which is an efficient way to group patients who recover similarly together. Concerning the other features that have not been used by the algorithm, Figure 4 gives a useful insight.

**Fig. 4.**
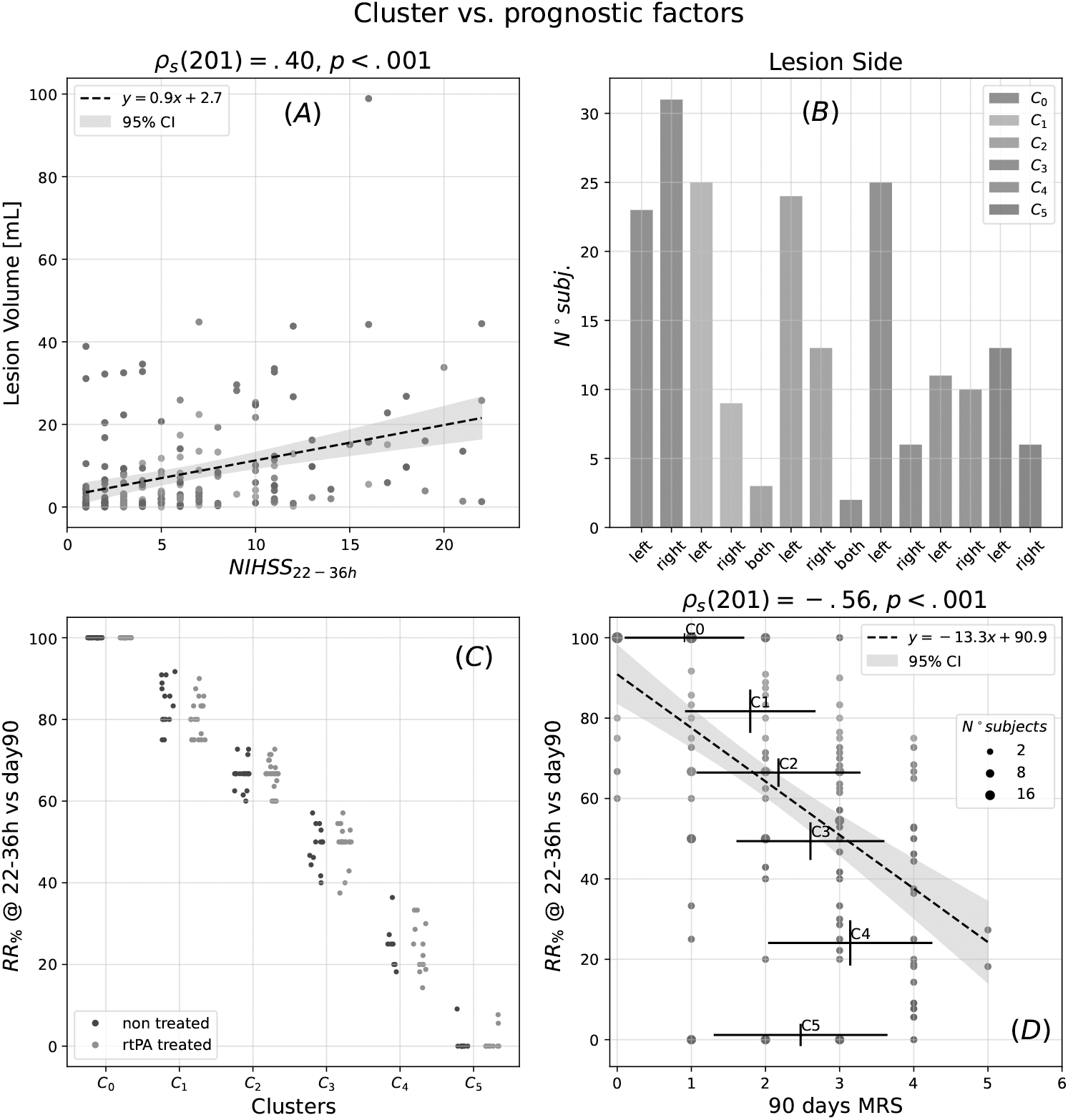
Cluster’s features. Panel A investigates the relation between lesion volume and total NIHSS at 22/36 hours. In both panels A and B, subjects (dots) are colored by cluster assignment, the dashed lines are the linear fitted-models, while the 95% confidence intervals (CI) are colored in gray. The lateralization distribution of the lesions (i.e. left, right or both) for each cluster is shown in panel B. Panel C shows the *RR* for the patients of each cluster, divided between rtPA treated group (magenta) and non-treated group (blue). Finally, Panel D shows instead the relation between the recovery ratio (*RR*) vs. 90 day MRS (modified rankin scale). Scatter size, represent the number of subjects having that value, as indicated by the corresponding legend in Panel D.

**Fig. 5.**
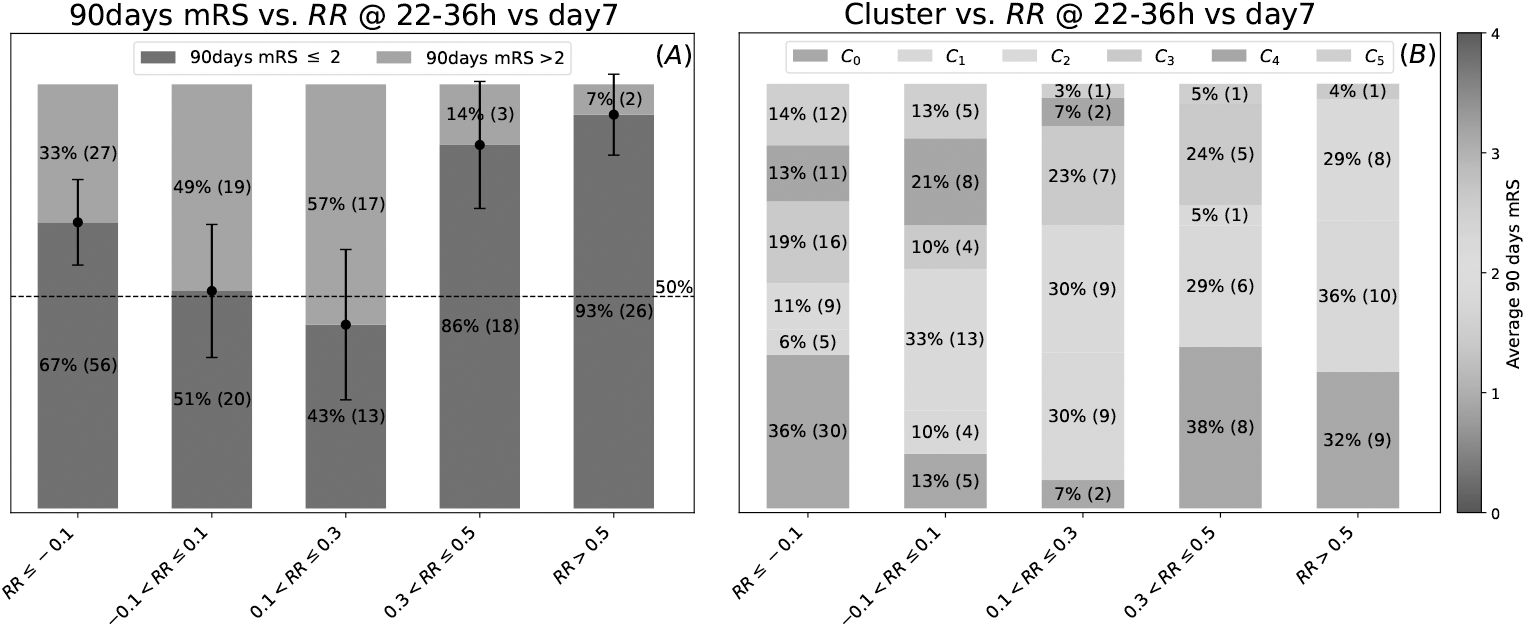
90days mRS relation with early recovery. Panel A shows the amount of patients for each range of recovery ratio, for each of the two range of the mRS considered. In detail in blue when mRS ≤ 2, while in orange *>* 2. 95% confidence interval are associated to each counts (errorbars), using the binomial distribution. The dashed line indicates the random chance of 50%. Panel B instead shows the number of subjects in each range of recovery ratio for each cluster. Each block corresponds to a cluster in descending order (*C*_0_ is always present on the bottom, in light blue). Color bar indicates the average 90 day mRS score for each cluster.

Panel A of Figure 4 shows the linear relation between the lesion volume (expressed in mL) and the sub-acute score; points are colored by cluster assignment. The two variables are significantly correlated (*ρ*_*s*_(201) = .40, *p <* .001), but there is no significant association between lesion volumes and clusters (Kruskal-Willis test, *H*(5) = 7, *p* = .22). In other words, there is no significant relation between lesion volume at admission and recovery measured at 90 days.

Panel B of Figure 4 represents the distribution of the lesion side (left, right, bilateral) in the obtained clusters. The clusters obtained with RSC show significant differences in lesion laterality (*χ*^2^(5, 201) = 25.6, *p* = .004, *V* = .25). In particular *C*_0_ includes more right lesions than expected, while *C*_3_ more left lesions (pairwisepost hoc test, *χ*^2^(1, 87) = 14.8, *p* = .009, *V* = .41). In other words, the sides of the lesions are not randomly divided by clusters. Subjects with perfect recovery (*C*_0_) have significantly more right than left lesions; conversely, patients in clusters with average recovery (*C*_3_) have significantly more left than right lesions.

In Figure 4 Panel C, the recovery ratio distribution is reported for each group, where patients have been separated between treated with rtPA (magenta) and non-treated (blue). As can be seen in Figure 4, there are no significant differences between the distributions of the two groups for each cluster, and there is no significant association between treatment and clusters (*χ*^2^(5, 201) = 5.7, *p* = .33). Therefore, while rtPA improves NIHSS and mRS scores acutely and at 3 months (see supplementary material Section 7, Figure 3), data show that patients treated with rtPA have the same recovery trajectories than patients non treated with rtPA after the first 24 hours from the event. Finally, no significant differences were found between the clusters with respect to the Heidelberg hemorrhagic classification.

The recovery ratio and the 90 days mRS are significantly correlated (*ρ*_*s*_(201) = .56, *p <* .001), and there is an association between the RSC clusters and motor abilities at 90 days (Kruskal-Willis’ test, *H*(5) = 74.3, *p <* .001, *η*^2^ = .36). Post hoc pairwise comparisons, conducted using the Mann-Whitney U test, reveal significant differences between *C*_0_ and all other clusters, *C*_1_ versus *C*_3_ and *C*_4_, and *C*_2_ versus *C*_4_. Finally, no cardiovascular risk factor is associated with a specific cluster, and we found no significant differences between clusters, both at *α* = 0.01.

### 3.3 Prediction of long-term recovery by early recovery ratio

In addition, we investigated whether cluster identity, as well as overall outcome of each patient was related to initial recovery. This analysis is particularly interesting from a clinical perspective and provides valuable prognostic insights for the patient and caregivers during hospitalization. We tested the relation between disability at 90 days and the early recovery ratio measured between 22/36 hours and 7 days, in relation to the identified clusters. The results are reported in Figure 5. In Panel A, it can be observed that a recovery ratio between −0.1 and 0.3 does not provide sufficient information to predict whether chronic disability will be severe or not (random chance level). In contrast, an early recovery ratio greater than 0.3 substantially increases the probability of achieving a mild disability. Panel B, shows the same result from a clustering point of view. A recovery ratio between −0.1 and 0.3 does not provide sufficient information to determine which recovery cluster the patient will belong to. The probability of being assigned to a group characterized by above average recovery (*C*_0_, *C*_1_ or *C*_2_) is 56% and 67% for the second and third ranges, and increases to 72% and 97% for the last two.

## 4 Discussion

In this work, we applied an unsupervised learning technique to the study of stroke recovery. Repeated spectral clustering (RSC), a recently developed procedure to tailor biomedical data heterogeneity [11], was applied to the recovery ratio based on total NIHSS in a large longitudinal sample of stroke patients. This approach grouped patients into distinct recovery clusters providing several clinical implications.

### 4.1 Recovery ratio and clustering

One of the main criticisms of the PRR is mathematical coupling, i.e. the fact that recovery is inherently dependent on the initial impairment. Essentially, from a mathematical point of view, fitting a linear model means establishing a linear relationship between an independent variable (the sub-acute score) and a dependent variable, the change (sub-acute score minus chronic score), which inherently includes the independent variable. This has caused much debate in the medical community, as evidenced by the extensive literature on the subject.

This methodology has been theoretically criticized by some and supported by others. The amount of recovery can be quantified as an absolute change, as previously mentioned, or in relative terms such as the recovery ratio index (*RR*). The definition of recovery ratio, in the framework of PRR, is induced by the variables chosen on the horizontal and vertical axes and by the linearity of the model. Instead, with clustering, the definition of recovery ratio could be carefully chosen a priori and not forced to be of the shape of Equation 1; the quantities of interest to quantify the recovery can be even nonlinearly correlated. Although it is true that *RR* defined by the PRR framework and used for this work has in its definition the ratio between chronic and acute scores, it is not calculated from a correlation or from the fit of a model; it simply represents a patient-specific estimate of recovery. In addition, this choice establishes a strong link between a traditional statistical framework and the clustering results obtained in this work.

Once the linear model is fitted, the identification of “fitters” vs. “non-fitters” may prove to be a great source of controversy. As shown in Figure 2, depending on the way patients are selected, different average recovery ratios can be estimated because different patients are included in the estimate. This shows how the same theoretical framework can give subjective outcomes. Clustering the recovery ratio instead is an effective way both to group patients with similar recovery and to incorporate time into a clustering algorithm. The six clusters found exhibit different levels of recovery, with patients described by their own PRR as indicated by Figure 3, Panel D. The prediction intervals of these models are narrow and well separate subjects into clusters. This clustering scheme may provide a novel methodology capable of fully exploiting longitudinal ordinal data, such as those presented in this paper.

### 4.2 Clinical implications

From a clinical perspective, the described dataset includes a large sample of stroke patients with an unknown time of onset. Wake-up strokes generally show poorer outcomes at three months than known onset strokes [29]. Although several studies have explored the relationship between this stroke category and revascularization therapies [30, 31], leading to progressively expanded treatment indications, research on general prognostic markers for recovery remains limited. In this context, we were able to provide a fine-grained description of this population over time. Specifically, in most patients, NIHSS scores progressively declined in a non-proportional manner (see from Panel D to Panel I, Figure 5 in the supplementary material, Section 8). In other words, most of the patients did not recover a fixed proportional amount of their initial deficit (see Figure 2). On the contrary, the identified clusters included patients with distinct trajectories, such as additive, proportional, or no improvement profiles, while still capturing a consistent amount of recovery during the first three months (i.e., the critical period for spontaneous recovery) [32]. This becomes particularly evident when further differentiating between severity categories for moderate and severe patient trajectories (see from Panel A to Panel C, Figure 5 in the supplementary material, Section 5).

The identified clusters are likely to reflect underlying neurophysiological differences that influence recovery trajectories. To evaluate this, we tested available prognostic factors (i.e. well-established post stroke recovery prognostic biomarkers) as validation features of the proposed approach. No significant differences in recovery ratio (*RR*) (i.e. from 22-36h to 90 days) were observed between patients who received rtPA and those who received a placebo. This finding is consistent with expectations, as *RR* calculation spans from 22–36 hours to 90 days post-stroke, whereas rtPA effects mainly affects the initial hours after the event [33]. Figures 3 and 4 in the supplementary material show improved early recovery (from onset to 22/36h) after acute revascularization treatment and no difference in the subsequent, spontaneous recovery from 22/36h to 90 days. A positive correlation between stroke volume and NIHSS at 22/36 hours supports the expected relationship between initial stroke severity and early impairment [34, 35]. However, stroke volume did not correlate with *RR*, indicating that lesion size alone does not predict long-term recovery patterns. This finding corroborates existing evidence that volume explains only a limited portion of outcomes, primarily related to initial impairment also following wake up stroke [36–38]. A negative correlation was observed between the mRS scores and both the *RR* and recovery groups, suggesting that patients with worse functional outcomes tend to exhibit poorer recovery trajectories. Worse recovery was also observed in patients with lesions of the left hemisphere. While this is consistent with some previous studies reporting worse functional outcomes for left-sided strokes in known-onset stroke [39], several studies have shown right hemisphere lesions also significantly affect recovery, especially reducing patients’ compliance to rehabilitation [40, 41]. Our results may, therefore, be explained by the NIHSS’s well-established increased sensitivity to left-sided strokes and should be validated using additional clinical scales in specific behavioral domains [42, 43]. Interestingly, no cluster presented a significantly higher frequency of patients with hemorrhagic transformation following rtPA. This suggests a relatively small impact of this feature on long-term disability of wake-up strokes undergoing thrombolytic treatment. These results underscore the substantial benefits of rtPA over its risks in this typology of stroke. Future work should evaluate the role of additional prognostic features of wake-up strokes, including vessel occlusion (i.e. anterior versus posterior circulation), comorbidites (i.e. hypertension as well as other cardiovascular risk factors) and neurophysiology indices (i.e. presence/absence of motor evoked potentials) on final disability, using a similar framework. Finally, patients who achieved an *RR* of 0.3 or higher within two weeks post-hospitalization consistently clustered into favorable outcome groups, with approximately 90% falling within clusters *C*_0_, *C*_1_, *C*_2_, and *C*_3_. A similar pattern emerged when considering mRS scores dichotomized for good (mRS ≤ 2) and poor (mRS *>* 2) outcomes. These results provide a valuable stratification, readily available during hospitalization (i.e. the first week after the event) with several prognostic implications for the patient and caregivers. In addition, these findings highlight the potential clinical application of this clustering approach, demonstrating its scalability across large datasets. Specifically, this methodology draws a useful parallel to how the mRS is employed in clinical trials for rtPA and thrombectomy. Just as these trials aim to identify an increased proportion of patients achieving favorable mRS outcomes, future interventions on neuroplasticity and functional re-organization after stroke could focus on shifting more patients toward the most favorable recovery clusters.

### 4.3 Limitations and future directions

In regards to the limitations of this study, a potential caveat of this approach is that for NIHSS at 22/36 hours, less than 6, the six linear models found with their relative prediction interval do not discriminate accurately between patients. This could somewhat limit the analysis by questioning the validity of these clusters and the separability between them. This is related to the “ceiling” effect. In fact, in the statistical model shown in Figure 3 Panel C, the prediction interval (in light blue) for a NIHSS less than 6 hardly discriminates points between “fitters” and “non-fitters”, questioning the validity of this subdivision for low-impaired subjects. Other clinical measures could be used to discriminate subjects even in this area of low impairment. Specifically, while the clustering approach was limited to only one variable (i.e. the recovery ratio), this was calculated from two variables at different times. Although the use of one variable could be criticized, it is worth underlining that clustering is not a dimensionality reduction technique. Future work could exploit the combination of additional variables for the purpose of phenotiping patients; nonetheless, clustering the recovery ratio can be seen as a clever strategy to embody time in a clustering algorithm. The full deficit dynamics at additional time points could be explored, enriching current knowledge about stroke recovery. Additional neurophysiological differences across clusters deserve further exploration. These include neuroimaging features (i.e. lesion location, lesion induced disconnection) as well as electro-physiological signals (i.e. evoked potentials and resting state electroencephalography). Finally, it should be noted that these results are limited to the sample in analysis and only indirectly account for several well described prognostic factors (i.e. hypertension, vessel occlusion, comorbidities, corticospinal tract integrity, among others). Therefore, replication in independent cohorts and evaluations using alternative clinical evaluations will be essential to validate and expand these results.

## 5 Conclusion

This paper aims to pave the way for a new, data-driven approach to studying stroke recovery. By using unsupervised techniques such as Repeated Spectral Clustering, we offer a more precise sub-division of patients based on their recovery trajectories. Clustering the recovery ratio is an efficient way to embody time within the algorithm, while also accommodating the inherent variability in recovery across individuals. This flexible and scalable framework supports the analysis of larger, more heterogeneous datasets and enables continuous evaluation of neurophysiological differences between groups. Ultimately, it offers insights into the mechanisms of post-stroke recovery that extend beyond the Proportional Recovery Rule.

## Data Availability

All data produced in the present study are available upon reasonable request to the authors

## 6 Acknowledgment

- Funding: This work was supported by the STARS@UNIPD funding program of the University of Padova, Italy, through the project: MEDMAX. This project has received funding from the European Union’s Horizon Europe research and innovation program under grant agreement no 101137074 - HEREDITARY. ALB, VV and MC were supported by HORIZON-ERC-SyG (Grant No.101071900) “Neurological Mechanisms of Injury And Sleep-Like Cellular Dynamics (NEMESIS)”. MC was supported by HORIZON-INFRA-2022 SERV (Grant No.101147319) “EBRAINS 2.0: A Research Infrastructure to Advance Neuroscience and Brain Health”.
- Conflict of interest: The authors have no competing interests to declare that are relevant to the content of this article.
- Code availability: code is available at MedMaxLab/beyondPRR.
- Authors’ contributions: AZ: Conceptualization, Data Curation, Methodology, Clustering, Statistical Analysis, Writing - Original Draft. ALB: Conceptualization, Methodology, Data Curation, Writing - Original Draft. VV: Methodology. MA: Project Administration, Supervision. MC: Conceptualization, Project Administration, Supervision. All authors reviewed and edited the manuscript.

