## Supplementary Material for "Beyond proportional recovery in wake-up stroke: unsupervised recovery clusters based on the NIHSS"

### 1 Supplementary Material

#### 1.1 Inclusion Criteria

The inclusion criteria followed to get the 201 patients from the WAKE-UP dataset [1] used for this work are schematized in Figure 1.

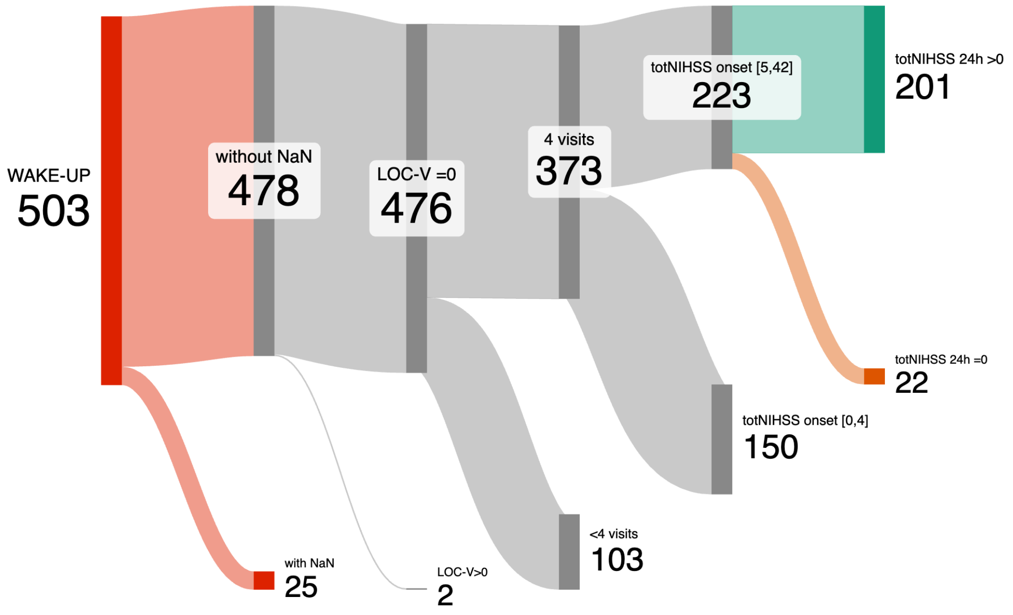


Figure 1: Sankey plot of patients selection used for this work. On the left side in red the whole WAKE-UP dataset (503 patients), on the right side in green the patients used in this work (201).

#### 1.2 PRR Model Formulation

In order to obtain the proportional recovery rule, a linear model of the type change -(chronic score - sub-acute score) can be fitted on the data. For clarity and limited to this section, let's define $y$ the chronic score and $x$ the sub-acute score. The linear model fitted on this work does not have the intercept, which is due to three reasons: the first is that, when a model with intercept is fitted, the coefficient takes positive value $(-(y-x) = mx + q, m = 0.49, q = 0.54)$. This means that when $x = 0$ (i.e no sub-acute impairment) we have $y < 0$ (i.e negative chronic impairment), which is impossible since the total NIHSS is a positive number by definition. Secondly, if the model is without the intercept, the slope $m$ is strictly related to the recovery ratio previously defined; indeed the definition of recovery ratio can be obtained by inverting the formula:

$$-(y-x) = mx \rightarrow -(y-x)/x = m, \ x \neq 0 \rightarrow 1 - y/x = m \tag{1}$$

However it is worth to underline that the slope $m$ and the average recovery ratio $\bar{RR}$ slightly differs in formulation, when estimated from the data in details

$$m = 1 - \frac{\sum_i x_i y_i}{\sum_i x_i^2}, \ \bar{RR} = 1 - \frac{1}{N}\sum_i \frac{y_i}{x_i} \tag{2}$$

thus the slope is $m = 0.54 \pm 0.02$, while the average recovery ratio for the whole population $\bar{RR}_{\text{negative}} = 0.58 \pm 0.67$ is slightly different. Finally, the $F$ test for nested models [2], turns out to be not significant ($F(1, 199) = 4.45,\ p = .036$), which favors the model with less parameters, i.e without the intercept.

#### 1.3 Statistics behind the PRR

In both panels of Figure 1 of the main manuscript, it has been calculated the 95% confidence interval (CI) as indicated in [3]; the CI describes the uncertainty over the fitted model and estimated parameters. We report in Equation 3 the analytical formula.

$$\hat{Y}_{ci} = \hat{Y} \pm t_{1-\alpha/2,n-2}\sqrt{\frac{\sum_i (y_i - \hat{y}_i)^2}{n-2}}\sqrt{\frac{1}{n} + \frac{X - \bar{x}}{\sum_i (x_i - \bar{x})}} \quad \alpha = 0.05,\ n = 201 \tag{3}$$

The prediction interval (PI) instead is an estimate of an interval in which a future observation will fall and it has been calculated as described in [3]. We report in Equation 4 the analytical formula.

$$\hat{Y}_{ci} = \hat{Y} \pm t_{1-\alpha/2,n-2}\sqrt{\frac{\sum_i (y_i - \hat{y}_i)^2}{n-2}}\sqrt{1 + \frac{1}{n} + \frac{X - \bar{x}}{\sum_i (x_i - \bar{x})}} \quad \alpha = 0.30,\ n = 201 \tag{4}$$

#### 1.4 Outliers identification

Eight different methods were tested to define "non-fitters" (outliers), and the average recovery ratios calculated for the "fitters" are reported in Figure 2 of the main manuscript. Below, we provide methodological details for each method.

1. No dichotomization. All subjects are considered "fitters", which gives an average recovery ratio of $RR_1 = 0.65 \pm 0.31$.
2. Outside the 70% prediction interval. Subjects outside the prediction interval (light blue band in Figure 1 of the main manuscript) are considered "non-fitters". This gives an average recovery ratio of $RR_2 = 0.69 \pm 0.27$.
3. Outside Percentile Range (a) $[\alpha; 1]$. Subjects with a recovery ratio outside this percentile range are considered "non-fitters". With $\alpha = 10\%$, this gives an average recovery ratio of $RR_3 = 0.72 \pm 0.25$.
4. Outside Percentile Range (b) $[\alpha/2; 1 - \alpha/2]$. Subjects with a recovery ratio outside this percentile range are considered "non-fitters". With $\alpha = 10\%$, this gives an average recovery ratio of $RR_4 = 0.63 \pm 0.31$.
5. Studentized Residuals. This method, used in [4], first selects patients showing a change above a certain threshold, then fits a linear model on the remaining data. After that, studentized residuals are computed, and subjects with $|t| > 2.31$ are classified as "non-fitters". Using $\Delta_{\text{NIHSS}} > 5$, this gives an average recovery ratio of $RR_5 = 0.66 \pm 0.30$.
6. Cumulative Correlation Coefficient. This method, used in [5], orders subjects based on initial impairment and calculates the Pearson correlation coefficient between initial score and change. This metric is computed iteratively by progressively adding more ordered data points. If, after a plateau, a deviation from the trend is observed, it could indicate the presence of "non-fitters", allowing for the arbitrary definition of a threshold to separate the two groups. Using $\text{NIHSS}_{\text{22-36h}} > 21$, this gives an average recovery ratio of $RR_6 = 0.65 \pm 0.31$.

7. Severely impaired patients. This method, used in [6] divides subjects based on initial impairment. Using $\text{NIHSS}_{\text{22-36h}} > 21$, this gives an average recovery ratio of $RR_7 = 0.65 \pm 0.32$.

8. Hierarchical clustering. This method, used in [7] applies agglomerative hierarchical clustering in the change vs. initial-score plane to cluster patients into two groups: "fitters" and "non-fitters". Average linkage with Mahalanobis distance was used, which gives an average recovery ratio of $RR_8 = 0.69 \pm 0.28$.

##### 1.5 Optimal $k$

The elbow-criteria was chosen as heuristic to find the optimal number of clusters; in details, the max-gap calculated for various $k$ is calculated and then the optimal $k$ is individuated as the highest $k$ with high max-gap [8]. The max-gap plot is reported in Figure 2.

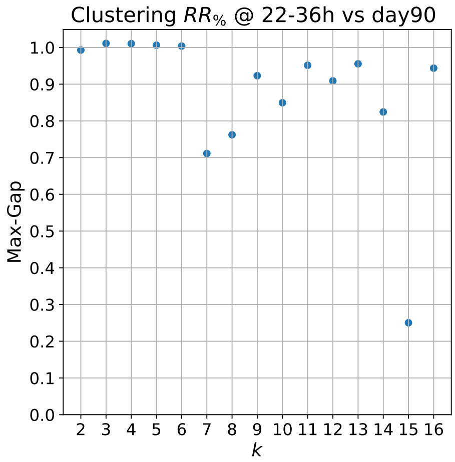


Figure 2: Max-gap calculation for various $k$. The optimal $k$ following the elbow-criteria is $k = 6$, which is highest $k$ with high max-gap.

##### 1.6 Effect size statistics

Cramer's $V$ [9] is used to quantify the effect size for the $\chi^2$ independence test [10].

$$V = \sqrt{\frac{\chi^2}{\min(k-1, r-1)N}} \tag{5}$$

Here $k$, $r$ are respectively the number of columns and rows, $N$ is the number of observations and $\chi^2$ the statistic of the test under analysis.
The eta-squared $\eta^2$ [11] is used to quantify the effect size for the Kruskal-Wallis' test [12].

$$\eta^2 = \frac{H - k + 1}{N - k} \tag{6}$$

Here $k$ is the number of groups, while $N$ is the number of observations and $H$ the statistic of the test under analysis.

The f-squared $f^2$ [13] is used to quantify the effect size for the $F$ test, for nested models.

$$f^2 = \frac{(RSS_1 - RSS_2)/(k_1 - k_2)}{RSS_1/(n - k_1)} \tag{7}$$

Here $RSS_1$ is the residual sum of squares for the more complex model, while $RSS_2$ for the simpler one; $k_1, k_2$ the number of parameters in each model, while $N$ is the number of observations.
The three effect size are then compared with the following grid, in order to interpret them [13].

| Effect Size | $V$ | $\eta^2$ | $f^2$ |
|---|---|---|---|
| Small | 0.1 | 0.06 | 0.10 |
| Medium | 0.3 | 0.15 | 0.30 |
| Large | 0.7 | 0.35 | 0.40 |

Table 1: Effect size and their interpretation.

#### 1.7 Treatment effect

The 201 subjects considered in this study are divided into two groups according to the treatment, specifically 94 treated and 107 non-treated.

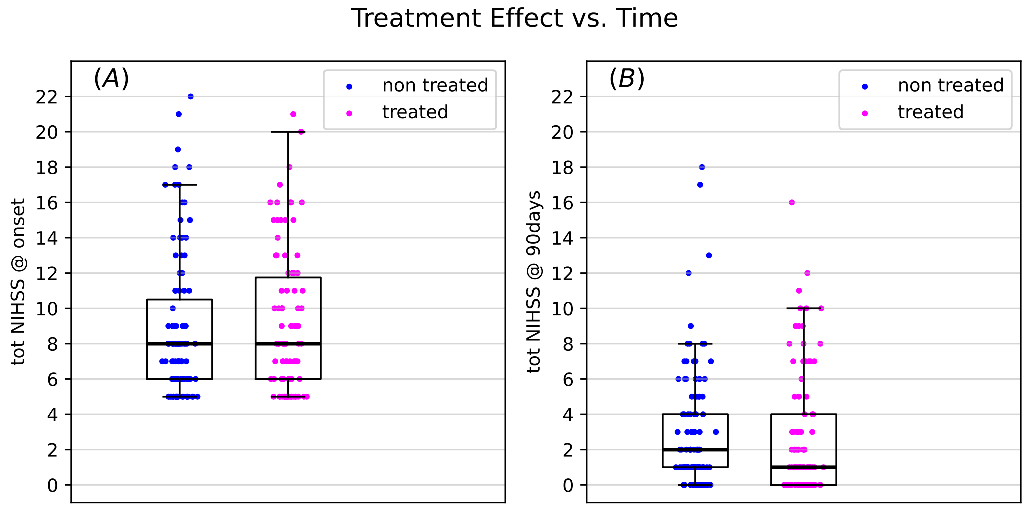


Figure 3: Effects of treatment on the NIHSS. The two groups treated (magenta) and non-treated (blue) are compared at onset (Panel A) and at 90 days (Panel B).

The NIHSS values of the two groups are recorded at the beginning of the study and after 90 days. In both cases, the four distributions were found to be non-normal ($p > 0.01$, Saphiro-Wilk's test), so a Mann–Whitney U test [14] was applied to compare the differences between the groups. No significant differences were found at $\alpha = 0.01$; however, at 90 days, a clear trend towards lower severity is evident in the treated group compared with the non-treated group. Analogously, the same can be done by inspecting the recovery ratios $RR$ calculated between different time points; while a clear trend is present, no significant results have been found.

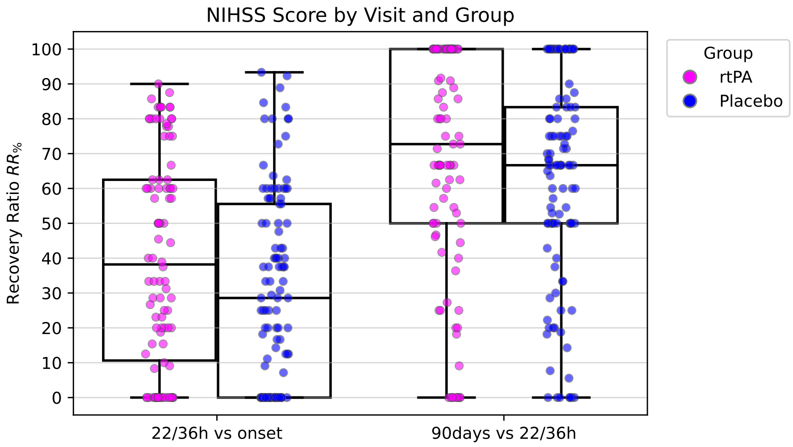


Figure 4: Effects of treatment on the recovery ratio ($RR$). The $RR$ of the two groups treated (magenta) and non-treated (blue) are compared to each other. $RR$ is calculated between onset and 22/36 hours on the left and between 22/36 hours and 90 days.

#### 1.8 Individual patient trajectories

For the 201 subjects considered in this study, the NIHSS trajectories for the four time points are reported in Figure 5. Subjects are divided by severity in Panels A, B and C, while they are divided by cluster from Panel D to Panel I.

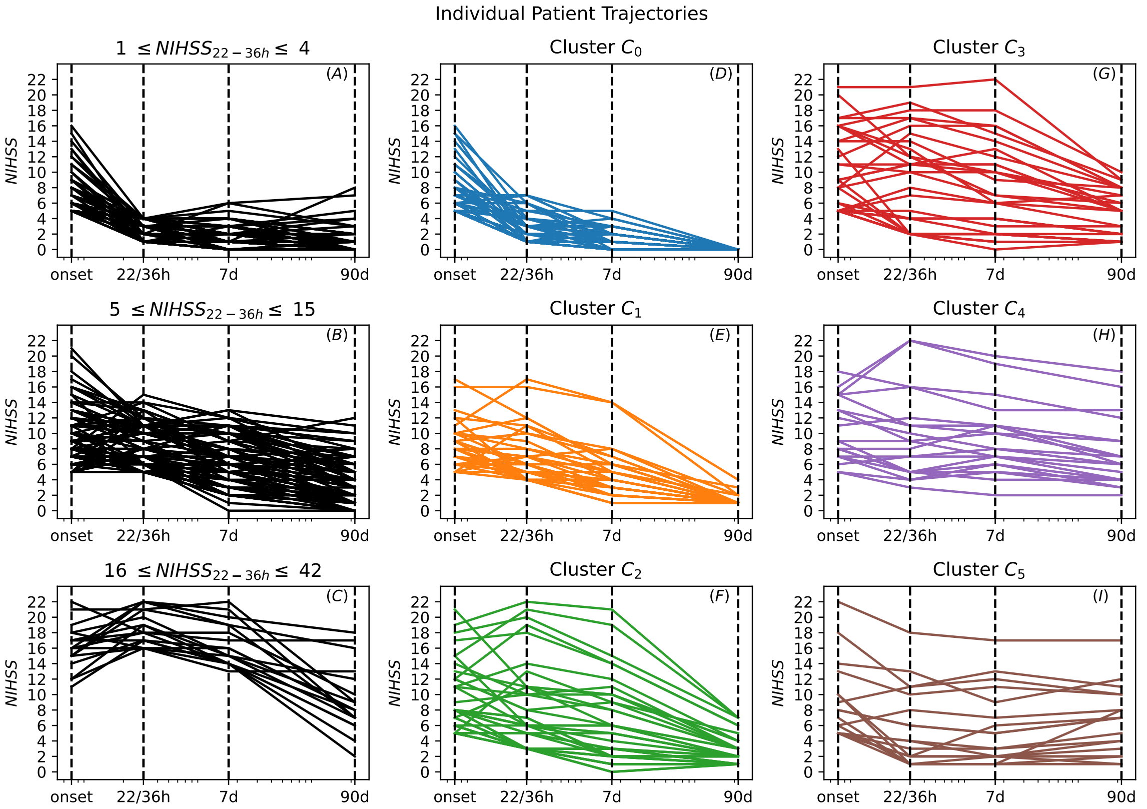


Figure 5: Individual patient trajectories. On the left side of the image, Panels A to C in black, there are the individual patient trajectories divided by severity measured at the sub-acute phase (22/36 hours). On the center-right side of the image, Panels D-I in colors, there are the individual patient trajectories divided by clusters assignment. On the x-axis, it is reported the time in logarithmic scale.
